# Ophthalmologic evidence against the interpersonal transmission of 2019 novel coronavirus through conjunctiva

**DOI:** 10.1101/2020.02.11.20021956

**Authors:** Yunyun Zhou, Yuyang Zeng, Yongqing Tong, Changzheng Chen

## Abstract

**Background:** The emerging 2019 novel coronavirus (2019-nCoV) has pushed several countries into state of emergency all over the world. The possible transmission of 2019-nCoV by conjunctiva is controversial and has substantial public health implications.

**Methods:** A retrospective cohort study was initiated to investigate the possible transmission of 2019-nCoV through aerosol contact with conjunctiva. We enrolled 67 cases of confirmed or suspected cases of novel coronavirus pneumonia (NCP) during 17–28 Jan 2020. Nasopharyngeal and conjunctival swabs were collected for real time RT-PCR analysis to detect 2019-nCoV.

**Results:** 63 patients were identified as laboratory-confirmed NCP and the remaining four were suspected NCP. Conjunctival swab samples from one NCP patient yielded positive PCR results and two NCP patients yielded probable positive PCR results. None of the three patients had ocular symptoms. The only one NCP patient with conjunctivitis as the first symptom had negative conjunctival sac 2019-nCoV test. Conjunctival swab samples from the four suspected cases of NCIP were negative.

**Conclusion:** 2019-nCoV can be detected in the conjunctival sac of patients with NCP. Through clinical analysis, viral transmission via the conjunctival route was not supported by the data. Good clinical protection can effectively cut off the transmission path.

## Introduction

The current outbreak of the 2019 novel coronavirus (2019-nCoV) originating in Wuhan, China has posed a threat to global public health and has been defined as a Public Health Emergency of International Concern (PHEIC) by the World Health Organization (WHO)^1^. Thus far, tens of thousands of confirmed cases, including healthcare workers, have been identified in China, with the latest mortality rate calculated at approximately 2.52%^2^. Generally, patients infected with 2019-nCoV develop respiratory illness, with the first symptoms of fever, cough and fatigue that quickly progress to pneumonia^3^. A number of patients were observed with extra-pulmonary manifestations at the onset of the illness, such as conjunctivitis, or even presented with asymptomatic infection^4,5^. Additionally, the susceptible and potential routes of viral spread from patients or asymptomatic carriers to healthy person have yet to be fully understood^6^. These issues pose great challenges for disease diagnosis and outbreak control, and make the goal of improving our understanding one of the upmost urgencies.

2019-nCoV has been identified as one of a class of single-stranded enveloped RNA viruses, belonging to the beta-coronaviruses genus of the *Coronaviridae* family that has been isolated from several mammals, including humans^7^. Although the genome sequence of 2019-nCoV is distinct from severe acute respiratory syndrome coronavirus (SARS-CoV) and Middle East respiratory syndrome coronavirus (MERS-CoV), they all have the potential to cause a severe acute respiratory disease and possess human-to-human transmissibility^8^. Similar to SARS-CoV, 2019-nCoV gains entry into host cells through recognizing and binding to its potential host receptor, angiotensin-converting enzyme 2 (ACE2), which is distributed among various tissue and cell types, including the conjunctiva^9-11^. A previous study indicates that healthcare workers suffer from higher risk of SARS infection in the case of unprotected eye contact with secretions^12^. The SARS-CoV has been shown to spread through direct or indirect contact with mucous membranes in the eyes^13,14^. Moreover, there are increasing reports suggesting that a few novel coronavirus pneumonia (NCP) cases began with conjunctivitis as the initial symptom following contact with confirmed patients without protective goggles^4,15,16^. For these reasons, determining whether 2019-nCoV is capable of transmission through aerosol contact with conjunctiva is an important consideration that warrants for further exploration.

In view of the limited understanding and controversy surrounding knowledge of viral transmission through the conjunctival sac, we performed a retrospective cohort study designed to seek ophthalmologic evidence for the transmission of 2019-nCoV from patients via aerosol contact with the conjunctiva. The findings of this study may provide novel insights into spread routes and potential infection control measures for 2019-nCoV that may help battle this outbreak.

## Methods

### Case enrollment

From 17 Jan to 28 Jan 2020, we enrolled 67 patients with either confirmed or suspected cases of NCP in the Renmin Hospital of Wuhan University. Laboratory-confirmed cases were identified with the criteria of at least one positive result from a respiratory specimen using viral isolation, next-generation sequencing, or reverse transcription-polymerase chain reaction (RT-PCR) assays. Suspected cases were recruited based on the criteria of fever or signs/symptoms of lower respiratory illness (e.g. cough or shortness of breath), low or normal white-cell count or low lymphocyte count, and no reduction in symptoms after antimicrobial treatment for 3 days. The history of epidemiologic exposure was also considered and incorporated^17^.

All patients invited to participate in the study provided consent for the nasopharyngeal and conjunctival swab samples. The study was approved by the Ethics Committees of Renmin Hospital of Wuhan University.

### Study design

Patient clinical details, including general information, clinical manifestations, and extra ocular symptoms, were obtained through review of health records or the implementation of questionnaire-styled interviews. General information was documented, including age, gender, underlying chronic conditions, date of symptom onset, date of hospital and ICU admission, and previous history of contact with confirmed patients or potential exposure to 2019-nCoV. The clinical manifestations included fever, cough, fatigue, shortness of breath, etc. Additionally, any ocular symptoms at the very onset, such as conjunctivitis or other ophthalmic manifestations, were solicited from all participants.

We took nasopharyngeal swabs and conjunctival swabs from all patients during hospitalization. Further laboratory methods were performed on these swab samples for the detection of 2019-nCoV utilizing real-time RT-PCR assays. The experimental protocols are as follows: RNA extraction from conjunctival swabs and nasopharyngeal swabs was performed in preparation for subsequent one-step reverse transcription–coupled PCR reaction using a proprietary master mix containing a DNA binding dye and thermal cycler. The primers and probes for 2019-nCoV detection were selected according to the National Pathogen Resource Center (National Institute for Viral Disease Control and Prevention, China CDC), and the sequences were as follows: forward primer 5′-CCCTGTGGGTTTTACACTTAA-3′; reverse primer 5′-ACGATTGTGCATCAGCTGA-3′; and the probe 5’-FAM-CCGTCTGCGGTATGTGGAAAGGTTATGG-BHQ1-3’. The conditions for the amplifications were 50°C for 15 min, 95°C for 3 min, followed by 45 cycles of 95°C for 15 s and 60°C for 30 s ^18^.

## Results

Samples were collected from 67 patients initially suspected to have NCP over a period of 11 days (17–28 Jan 2020), of whom, 25 were male and 42 were female (Table 1). The mean age was 35.7 (SD 10.6) years (range 22–78 years). The majority were healthcare workers and female nurses. By Jan 28, 2020, 63 patients were identified as laboratory-confirmed NCP and the other four were suspected NCP. Of the four suspected NCP cases, there were two males and two females. Conjunctival swab samples from one NCP patient yielded positive PCR results, and swabs from two NCP patients yielded probable positive PCR results. None of the three patients had ocular symptoms. The only one NCP patient with conjunctivitis as the first symptom had negative conjunctival sac 2019-nCoV test. Conjunctival swabs samples from the four suspected NCP cases were negative. The history of one positive case, two probable positive cases, and one case with the first symptom of the eye were as follows:

**Table 1.**
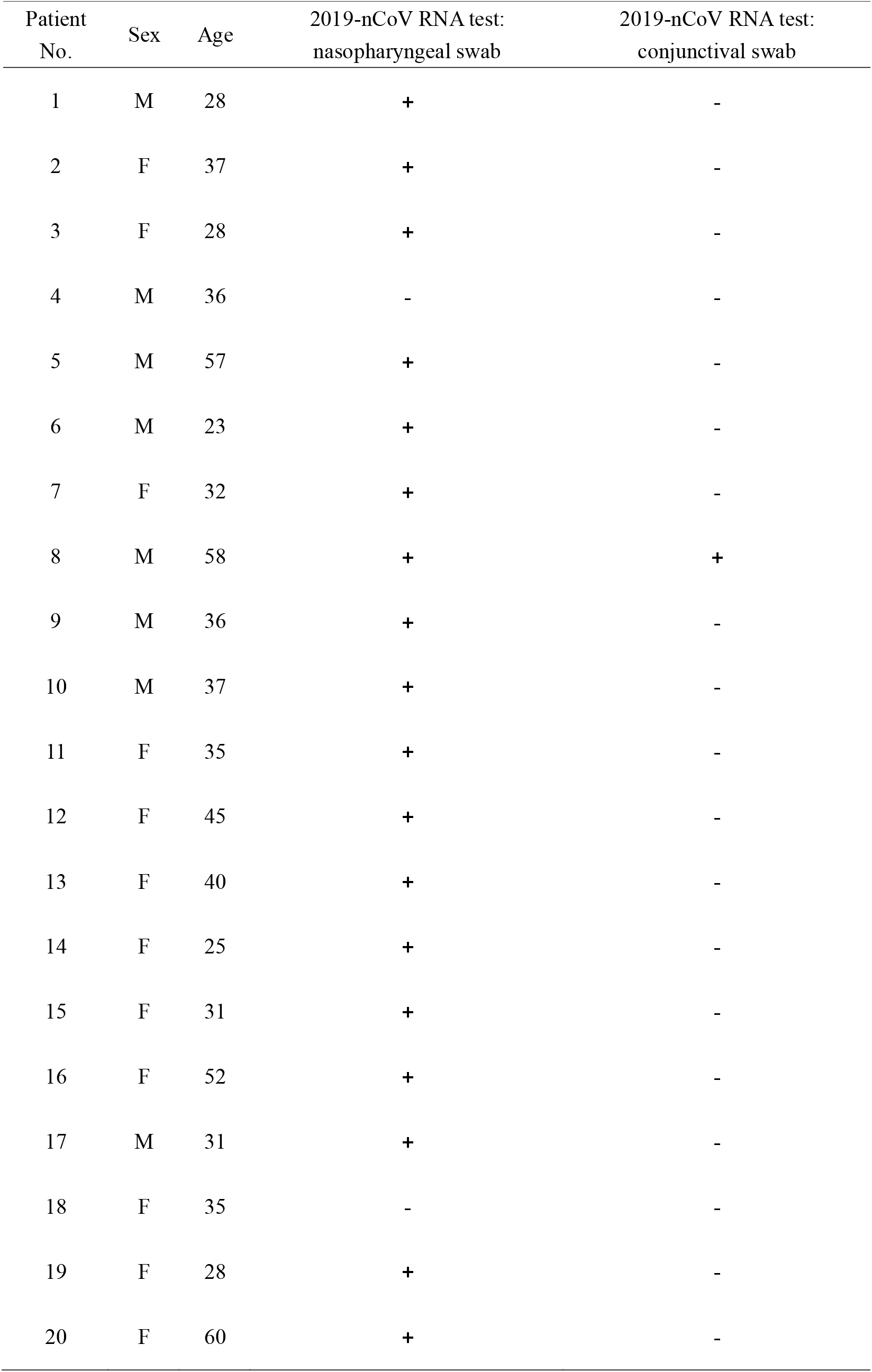
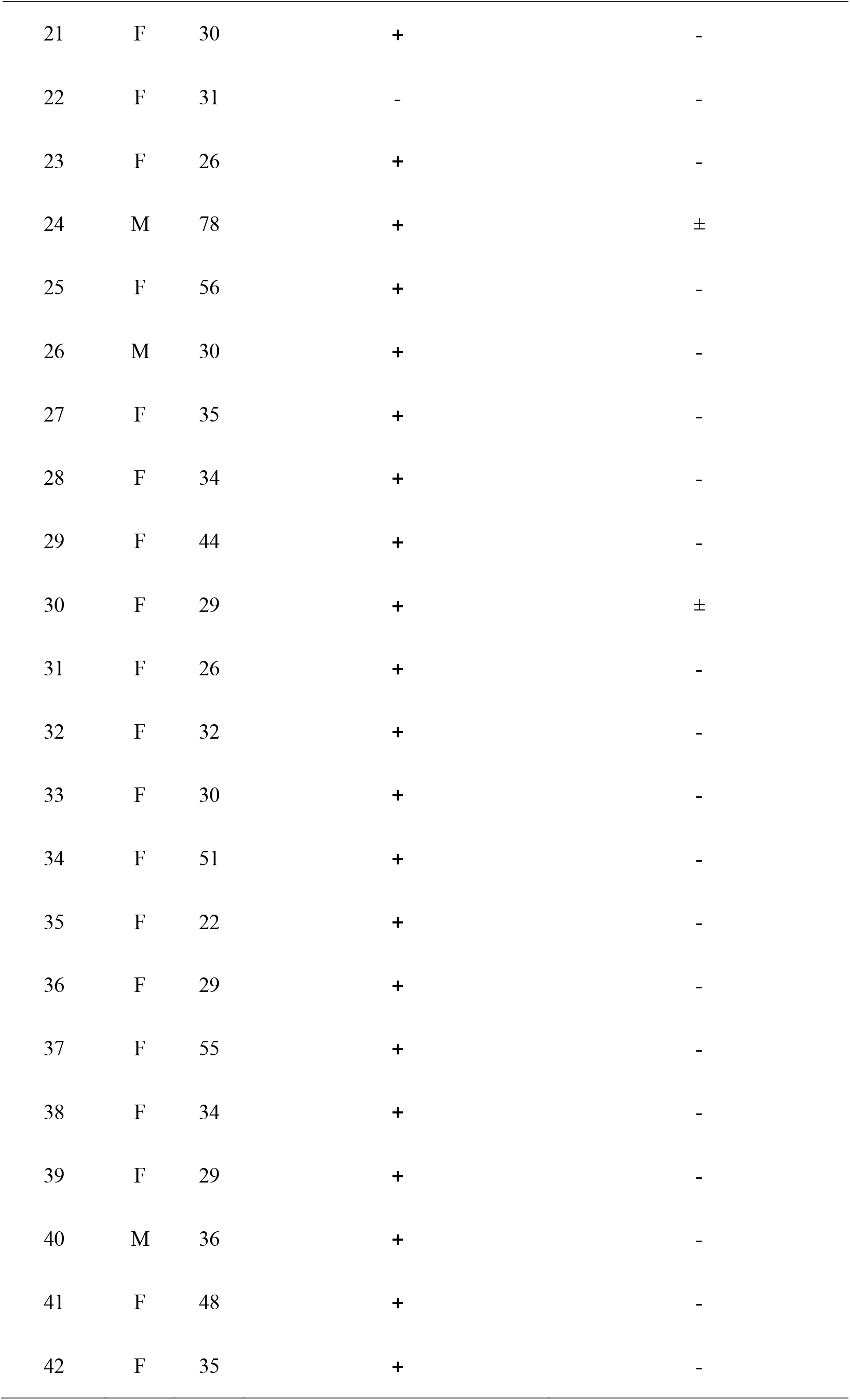
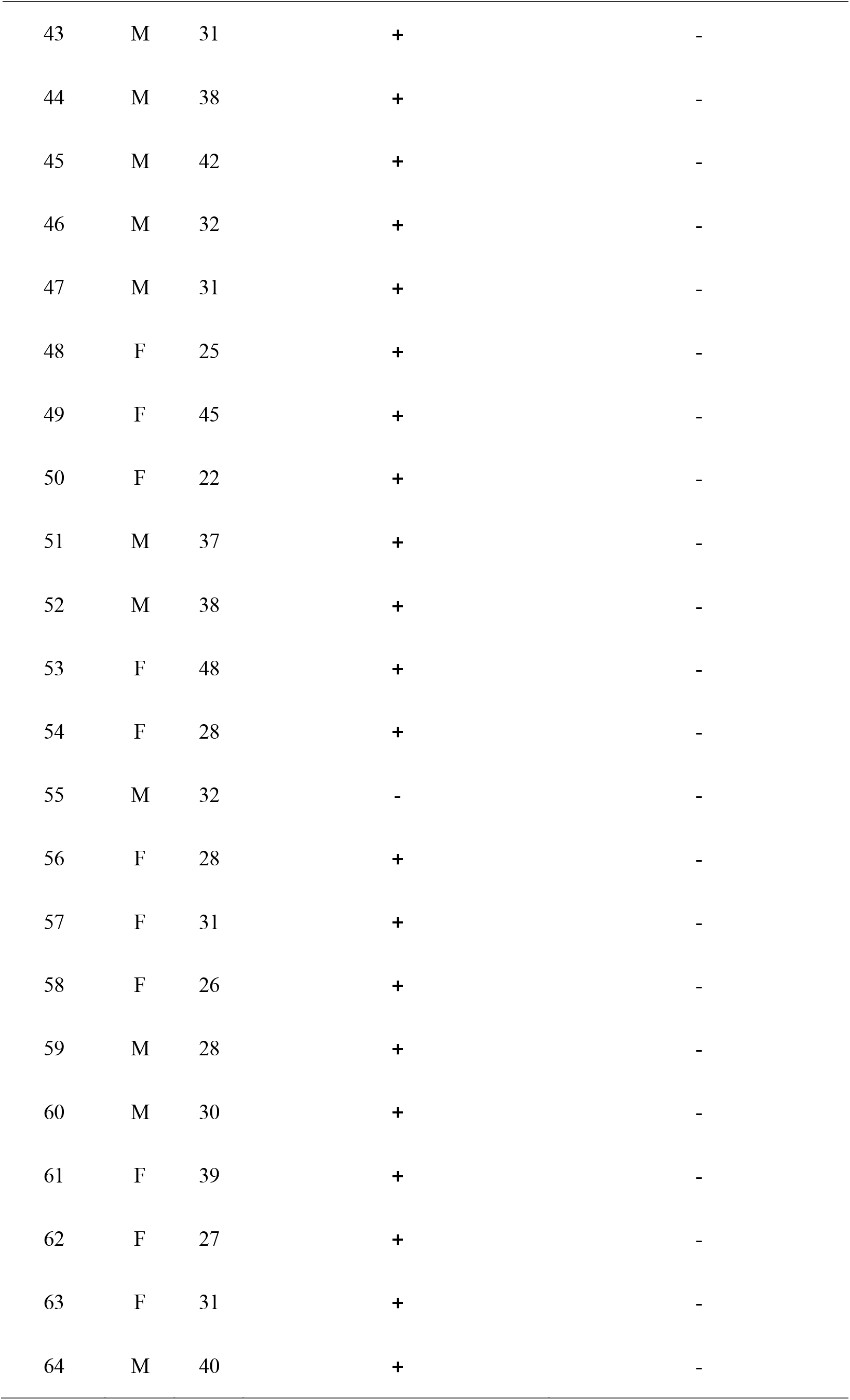
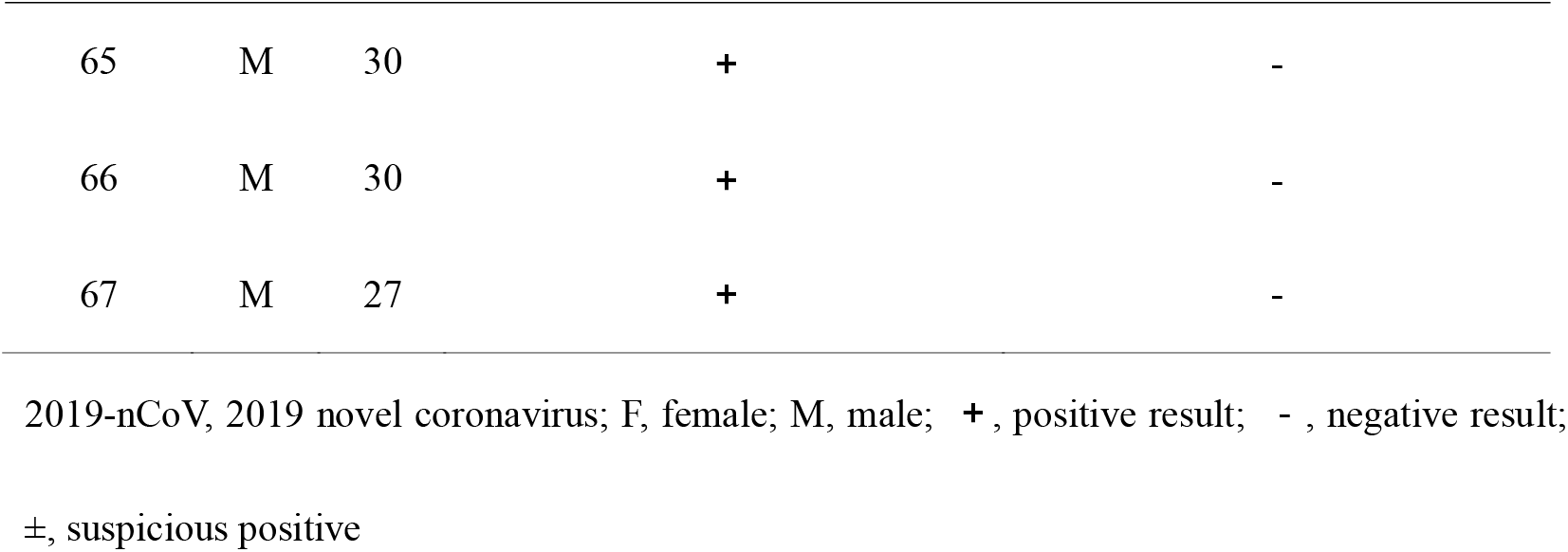
Signalment for 67 patients and viral detection results of nasopharyngeal and conjunctival samples.

| Patient No. | Sex | Age | 2019-nCoV RNA test:<br>nasopharyngeal swab | 2019-nCoV RNA test:<br>conjunctival swab |
| --- | --- | --- | --- | --- |
| 1 | M | 28 | + | - |
| 2 | F | 37 | + | - |
| 3 | F | 28 | + | - |
| 4 | M | 36 | - | - |
| 5 | M | 57 | + | - |
| 6 | M | 23 | + | - |
| 7 | F | 32 | + | - |
| 8 | M | 58 | + | + |
| 9 | M | 36 | + | - |
| 10 | M | 37 | + | - |
| 11 | F | 35 | + | - |
| 12 | F | 45 | + | - |
| 13 | F | 40 | + | - |
| 14 | F | 25 | + | - |
| 15 | F | 31 | + | - |
| 16 | F | 52 | + | - |
| 17 | M | 31 | + | - |
| 18 | F | 35 | - | - |
| 19 | F | 28 | + | - |
| 20 | F | 60 | + | - |
| 21 | F | 30 | + | - |
| 22 | F | 31 | - | - |
| 23 | F | 26 | + | - |
| 24 | M | 78 | + | ± |
| 25 | F | 56 | + | - |
| 26 | M | 30 | + | - |
| 27 | F | 35 | + | - |
| 28 | F | 34 | + | - |
| 29 | F | 44 | + | - |
| 30 | F | 29 | + | ± |
| 31 | F | 26 | + | - |
| 32 | F | 32 | + | - |
| 33 | F | 30 | + | - |
| 34 | F | 51 | + | - |
| 35 | F | 22 | + | - |
| 36 | F | 29 | + | - |
| 37 | F | 55 | + | - |
| 38 | F | 34 | + | - |
| 39 | F | 29 | + | - |
| 40 | M | 36 | + | - |
| 41 | F | 48 | + | - |
| 42 | F | 35 | + | - |
| 43 | M | 31 | + | - |
| 44 | M | 38 | + | - |
| 45 | M | 42 | + | - |
| 46 | M | 32 | + | - |
| 47 | M | 31 | + | - |
| 48 | F | 25 | + | - |
| 49 | F | 45 | + | - |
| 50 | F | 22 | + | - |
| 51 | M | 37 | + | - |
| 52 | M | 38 | + | - |
| 53 | F | 48 | + | - |
| 54 | F | 28 | + | - |
| 55 | M | 32 | - | - |
| 56 | F | 28 | + | - |
| 57 | F | 31 | + | - |
| 58 | F | 26 | + | - |
| 59 | M | 28 | + | - |
| 60 | M | 30 | + | - |
| 61 | F | 39 | + | - |
| 62 | F | 27 | + | - |
| 63 | F | 31 | + | - |
| 64 | M | 40 | + | - |
| 65 | M | 30 | + | - |
| 66 | M | 30 | + | - |
| 67 | M | 27 | + | - |

2019-nCoV, 2019 novel coronavirus; F, female; M, male; +, positive result; -, negative result;
±, suspicious positive

Patient No. 8 was a male, 58 years old, and an emergency vehicle driver for the pre-hospital emergency team. He was admitted to the hospital because of fever and cough. The chest CT examination showed viral pneumonia. Both the nasopharyngeal and conjunctival swabs tested positive for 2019-nCoV nucleic acid. There has been no ocular discomfort. He had a history of unprotected contact with isolated patients.

Patient No. 24 was a male, 78 years old, and had a history of lung cancer with brain metastasis, chronic obstructive pneumonia, and hypertension. He was admitted to hospital for coughing, expectoration, and gasping. The results of 2019-nCoV RNA testing revealed that the nasopharyngeal swab was positive and the conjunctival swab was suspicious positive. There was no ocular discomfort. He had a history of contact with an NCP recessive carrier. The recessive carrier was also diagnosed with NCP and was hospitalized.

Patient No. 30 was a female, 29 years old, and pregnant at 36 weeks. She was a physician. She was admitted to the hospital because of fever. The results of the 2019-nCoV RNA test revealed that the nasopharyngeal swab was positive and the conjunctival swab was suspicious positive. She had no ocular complaint. She had a history of contact with a confirmed NCP patient.

Patient No. 41 was a female, 48 years old, and an anesthesiologist. She was admitted to hospital with fever, cough, redness of eyes, itching, and secretion. The results of 2019-nCoV RNA testing revealed that the nasopharyngeal swab was positive but the conjunctival swab was negative. The anesthesiologist presented with conjunctivitis as the initial symptom. The ocular symptoms were mild. The discomfort of the eye was relieved by itself without medication. She had a history of contact with a confirmed NCP patient wearing a surgical mask but no protective goggles.

## Discussion

Of the cases enrolled in this study, one was positive for the conjunctival sac 2019-nCoV test and two cases were suspicious positive. None of these three patients had ocular symptoms. One anesthesiologist presented with conjunctivitis as the first symptom but had a negative conjunctival sac 2019-nCoV test. There have been some reports regarding conjunctival sac and coronavirus, and coronavirus has been isolated from 7-month-old children with conjunctivitis and bronchitis^19^. Moreover, SARS coronavirus was detected in the tears of SARS patients, although the detection rate of coronavirus in the conjunctival sac was low^20^. The results of this study revealed that the incidence of conjunctivitis in patients with NCP is not high. At present, this kind of conjunctivitis has no specific manifestation, and can present in one eye or two eyes. In the early stage, it appears as common conjunctival hyperemia with fewer secretions. It is watery and akin to thin mucus. Occasionally small pieces of conjunctival hemorrhage are seen. The ocular symptoms of the patients were mild and tended to be self-healing. There was great variation between the patients.

Our confirmed patient No. 41 was an anesthesiologist. She developed ocular symptoms after performing intubation anesthesia for a patient, followed by fever and cough. The patient was diagnosed with NCP, however during the anesthesia, the anesthesiologist wore only an ordinary surgical mask, hats, and gloves, and did not wear goggles, protective clothing, or other protective devices. Two surgeons (patient No. 10 and patient No. 40) who operated on the NCP patient were later diagnosed with NCP. The two surgeons had no ocular discomfort. General anesthesia involves tracheal intubation and may increase the risk of viral infection. Our confirmed patient No. 30 is a physician. She and her three colleagues (patient No. 28, patient No. 46, and patient No. 65) were all infected from the same patient. The conjunctival swab tests for the six infected doctors (patient No. 41, patient No. 10, patient No. 40, patient No. 28, patient No. 46, and patient No. 65) were all negative, which did not support the spread of the virus through temporary aerosol contact with the conjunctiva.

This is a retrospective study of a small sample size, in which there is only one time point. Negative results from conjunctival sac swabs could be influenced by the sampling amount and time, as viral conjunctivitis is self-healing. There was a certain false negative rate in the viral RNA test, and with this knowledge, we will continue to pay close attention to the progress of the patients. Additionally, we should determine whether 2019-nCoV is also present in convalescent patients as well as the infectivity of the conjunctiva.

Our clinical observation and analysis can serve as a reminder that all medical workers, not only ophthalmologists, must make solid efforts for protection when treating patients. Specifically, doctors must wear masks, goggles, protective clothing, and gloves. Following contact with patients, close attention must be paid to hand disinfection as well as the disinfection of relevant inspection instruments and the clinical environment. The above measures can serve to cut off transmission channels and prevent cross infection in support of public health and safety.

In conclusion, 2019-nCoV can be detected in the conjunctival sac of patients with NCP. Through our clinical analysis, the concept that the virus is transmitted through the conjunctival route it is not supported. The prevention of and treatment for new coronavirus infectious disease requires greater attention and activity. The possibility of eye infection and the ocular route as a potential infection source should be considered and further examined, and scientific protection should be carried out.

## Data Availability

all data generated or used during this study are available from the corresponding author by request.

## Acknowledgement

The authors wish to thank the library of Wuhan University and the help of the healthcare workers from Renmin Hospital of Wuhan University.

## References

1. Mahase E. China coronavirus: WHO declares international emergency as death toll exceeds 200. BMJ 2020;368:m408.

2. National Health Commission of the People’s Republic of China. Pneumonia epidemic situation of new coronavirus infection on February 9, 2020. 2020. (Accessed February 10, 2020, at http://www.nhc.gov.cn/xcs/yqfkdt/202002/167a0e01b2d24274b03b2ca961107929.shtml.)

3. Huang C, Wang Y, Li X, et al. Clinical features of patients infected with 2019 novel coronavirus in Wuhan, China. Lancet (London, England) 2020.

4. American Academy of Ophthalmology. Alert: Important coronavirus context for ophthalmologists. 2020. (Accessed January 28, 2020, at https://www.aao.org/headline/alert-important-coronavirus-context.)

5. Rothe C, Schunk M, Sothmann P, et al. Transmission of 2019-nCoV Infection from an Asymptomatic Contact in Germany. N Engl J Med 2020.

6. Zhang H, Kang Z, Gong H, et al. The digestive system is a potential route of 2019-nCov infection: a bioinformatics analysis based on single-cell transcriptomes. bioRxiv 2020:2020.01.30.927806.

7. Zhu N, Zhang D, Wang W, et al. A Novel Coronavirus from Patients with Pneumonia in China, 2019. N Engl J Med 2020.

8. Benvenuto D, Giovannetti M, Ciccozzi A, Spoto S, Angeletti S, Ciccozzi M. The 2019-new coronavirus epidemic: evidence for virus evolution. J Med Virol 2020.

9. Wan Y, Shang J, Graham R, Baric RS, Li F. Receptor recognition by novel coronavirus from Wuhan: An analysis based on decade-long structural studies of SARS. J Virol 2020.

10. Hamming I, Timens W, Bulthuis ML, Lely AT, Navis G, van Goor H. Tissue distribution of ACE2 protein, the functional receptor for SARS coronavirus. A first step in understanding SARS pathogenesis. J Pathol 2004;203:631–7.

11. Holappa M, Vapaatalo H, Vaajanen A. Many Faces of Renin-angiotensin System - Focus on Eye. Open Ophthalmol J 2017;11:122–42.

12. Raboud J, Shigayeva A, McGeer A, et al. Risk factors for SARS transmission from patients requiring intubation: a multicentre investigation in Toronto, Canada. PLoS ONE 2010;5:e10717.

13. Peiris JSM, Yuen KY, Osterhaus ADME, Stöhr K. The severe acute respiratory syndrome. N Engl J Med 2003;349:2431–41.

14. Belser JA, Rota PA, Tumpey TM. Ocular tropism of respiratory viruses. Microbiol Mol Biol Rev 2013;77:144–56.

15. Peking University Hospital Wang Guangfa disclosed treatment status on Weibo and suspected infection without wearing goggles. 2020. (Accessed February 9, 2020, at http://www.bjnews.com.cn/news/2020/01/23/678189.html.)

16. Lu C-w, Liu X-f, Jia Z-f. 2019-nCoV transmission through the ocular surface must not be ignored. The Lancet.

17. Li Q, Guan X, Wu P, et al. Early Transmission Dynamics in Wuhan, China, of Novel Coronavirus-Infected Pneumonia. N Engl J Med 2020.

18. Institute of Virus Disease Control and Prevention, China CDC. Primers and probes for detection of the novel coronavirus. 2020. (Accessed February 3, 2020, at http://ivdc.chinacdc.cn/kyjz/202001/t20200121_211337.html.)

19. van der Hoek L, Pyrc K, Jebbink MF, et al. Identification of a new human coronavirus. Nat Med 2004;10:368–73.

20. Loon SC, Teoh SC, Oon LL, et al. The severe acute respiratory syndrome coronavirus in tears. The British journal of ophthalmology 2004;88:861–3.

